# A Behavioral Economic Analysis of Demand for Health Insurance in Nigeria

**DOI:** 10.1101/2025.08.27.25334236

**Authors:** Promise Tewogbola, Deborah Odaudu, Oluwakamikun Adekunle, Tolulope Oyewole, Victoria Bodunde, Alan Franklin, Natalie Buddiga, Olaniyi Ige, Ejura Salihu, Justin T. McDaniel

## Abstract

Health insurance coverage remains critically low in developing economies, with Nigeria’s National Health Insurance Scheme covering only 5% of the population. Understanding consumer demand patterns is essential for designing effective insurance policies, yet limited research has applied behavioral economic frameworks to health insurance decision-making in developing contexts. This mixed-methods study investigated factors influencing health insurance demand in Nigeria using behavioral economic principles, examining the effects of market structure (open vs. closed economy), demographic characteristics, and price sensitivity patterns on insurance purchasing decisions. Study 1 employed a hypothetical purchase task with 76 Nigerian participants randomly assigned to open- or closed-economy conditions, measuring willingness to purchase health insurance at 15 price points (₦5–₦560000). Exponential demand curves were fitted using a modified exponential demand model to assess demand intensity (*Q*_0_) and price sensitivity (*α*). Study 2 involved focus group discussions with 5 participants who completed the purchase task, exploring decision-making processes through thematic analysis. The exponential demand model demonstrated strong predictive validity (median *R*^2^ = 0.87, IQR = 0.81–0.92) across demographic subgroups. Contrary to theoretical expectations, no significant differences emerged between open and closed economy conditions (*p >* 0.05). Age, income, gender, and current insurance status were significantly associated with demand parameters. Younger participants showed higher demand intensity but greater price sensitivity, while low-income participants demonstrated higher demand intensity (*Q*_0_ = 91.37) than high-income participants (*Q*_0_ = 74.29). Qualitative findings revealed “reverse price sensitivity,” where participants rejected very low-priced options due to quality concerns, and identified a potential pricing corridor of ₦3,000–₦17,500 ($2–$12 USD) balancing affordability with quality signaling. Behavioral economic models may effectively characterize health insurance demand patterns in Nigeria, revealing complex price-quality relationships that challenge traditional economic assumptions. The null finding for market structure effects suggests that existing healthcare alternatives and institutional distrust may override theoretical market distinctions. Results support targeted subsidies for younger and lower-income populations and highlight the importance of pricing strategies that signal quality while maintaining affordability.

## 1 Introduction

Health insurance represents a mechanism through which individuals transform unpredictable health risk into manageable financial obligations. Through risk pooling, individuals contribute to a collective fund that absorbs the financial impact of unexpected medical expenses, preserving personal and familial economic stability when health crises arise (Erlangga et al., 2019; Iqlima, 2024). Beyond this protective function, health insurance fundamentally also alters healthcare engagement patterns by subsidizing the costs of routine screenings, vaccinations, and early interventions. This preemptive access reduces the severity and cost of future health issues while also incentivizing proactive healthcare utilization, effectively shifting the decision calculus from reactive care to strategic, long-term well-being (Finkelstein et al., 2012; Dey & Bach, 2019).

Despite the critical role of health insurance, substantial coverage gaps persist globally, with particularly acute challenges in developing economies. Multiple interconnected barriers discourage enrollment: financial constraints render premiums unaffordable for many households, limited public awareness reduces understanding of insurance benefits, and weak institutional trust undermines confidence in insurance providers (Akinyemi et al., 2021; Ajike et al., 2020; Amu et al., 2022; Eze et al., 2024). Compounding these issues, individuals often underestimate future health risks, particularly when current health status is perceived as good (Kunreuther & Pauly, 2015). Coverage disparities are most pronounced in the absence of employer or government subsidies. In Nigeria, for example, the formal sector, primarily urban and salaried workers, constitutes most enrollees, while the informal sector, concentrated in rural areas, faces barriers of cost, knowledge, and access, leading to exclusion from prepayment schemes and continued reliance on out-of-pocket payments (Ajike et al., 2020).

Developing economies face unique challenges that significantly impact their healthcare systems and insurance markets. Healthcare systems in Africa often suffer from neglect and underfunding, leading to severe challenges across the six World Health Organization (WHO) pillars of healthcare systems, with man-made issues affecting service delivery, healthcare workforce, healthcare information systems, medicines and technologies, financing, and leadership/governance (Oleribe et al., 2019). Nigeria exemplifies these challenges, with its developing economy facing a myriad of challenges including economic stagnation, social inequalities, and inadequate infrastructure that collectively constrain access to essential healthcare services (Adams, 2024). Despite the goal of Nigeria’s National Health Insurance Scheme (NHIS) to improve coverage, enrollment remains low, with only about 5% of the population registered as of 2022 (Nigerian Institute of Social and Economic Research, 2024). Affordability remains a key barrier, compounded by reliance on out-of-pocket (OOP) payments, covering approximately 75% of healthcare expenditures, and limited public investment in health insurance programs (Abubakar et al., 2022; Aron-Dine et al., 2013; PwC, 2024; Sanil & Eminer, 2021). Other major issues include inefficient service delivery, inadequate healthcare infrastructure, and poor resource management, leading to substandard care quality (Eze et al., 2024). These challenges result in delayed care, high financial burdens, and unmet healthcare needs, with all the attendant adverse impact on quality of care, outcomes, cost and quality of life, particularly in rural areas (Abah, 2023).

The dynamics of health insurance participation are influenced by principles of supply and demand, shaped by the structure of the market itself (Reed et al., 2013). In a closed economy, the reinforcer, in this case, healthcare, is accessible solely through one channel, such as employer-sponsored or private insurance, requiring individuals to “work” (e.g., pay premiums) for every unit of access (Gilroy et al., 2018). This structure isolates the relationship between cost and consumption, often resulting in inelastic demand, where rising costs do little to deter use because alternatives are absent (Hursh, 2014). In contrast, an open economy provides multiple pathways to access healthcare, such as alternative providers, informal networks, or community schemes, resulting in more elastic demand. Here, consumers can substitute or forego formal insurance if costs become prohibitive. Open economies reflect real-world conditions where access is rarely confined to a single source, whereas closed systems, though rare outside experimental settings, offer clarity in isolating consumption dynamics (Gilroy et al., 2018). Yet, despite its relevance, empirical research on how open versus closed structures influence insurance demand, especially in developing economies such as Nigeria, is scarce. Understanding these dynamics is vital for policymakers, as market design profoundly impacts accessibility, affordability, and health outcomes (Aljuiaid et al., 2022; Hussain et al., 2024; Schoen et al., 2010).

Behavioral economics provides a valuable framework for understanding health insurance decision-making by integrating insights from psychology into economic decision-making (Hursh et al., 2020). Within this framework, demand is conceptualized as the degree to which some organism work in order to access some level of consumption of a reinforcer (Gilroy et al., 2020). As a result, a decision-maker is likely to invest more effort for commodities they value highly and less for those of lower value. The relationship between the quantity consumed of a commodity and its unit price can be modeled by a nonlinear, positively decelerating curve (Allen, 1938; Jacobs & Bickel, 1999). The model is represented by the following equation by Koffarnus et al. (2015):

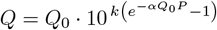

where *Q* is the quantity of the commodity consumed, and *Q*_0_ represents the highest level of demand when that commodity is free. The parameter *α* denotes the sensitivity of consumption to increases in cost. *P* stands for price, and *k* is the scaling constant reflecting the range of consumption data in log units.

Understanding consumer behavior is crucial for developing effective health insurance policies in developing economies such as Nigeria (Anyadighibe, 2022), where conventional research methods to measure health insurance demand may face logistical and ethical challenges (Adeleye & Adebamowo, 2012; Chukwu et al., 2016). Among behavior economic methodologies, Hypothetical Purchase Tasks (HPTs) offer a valuable methodological alternative, enabling researchers to model demand in a cost- and time-effective manner while also circumventing the ethical and practical limitations such as exposing participants to financial risk or psychological distress (Reed et al., 2022). HPTs have been validated across diverse commodities and populations, with successful applications in modeling demand for condoms (Strickland et al., 2020), gym memberships (Brown et al., 2021), COVID-19 vaccines (Hursh et al., 2020) and HIV vaccines (Tewogbola et al., 2025). Despite concerns about their hypothetical nature, HPTs reliably show consistent correspondence between stated preferences and observed market behavior (Khan, 2020; Murphy & MacKillop, 2006; Secades-Villa, et. Al., 2016; Strickland et al., 2022) and their flexibility makes them well-suited for contexts where large-scale trials are impractical or ethically fraught (Berry et al., 2023).

Consequently, the present study has three aims: (1) To evaluate the utility of behavioral economic demand curves, generated through HPTs, in describing health insurance demand in developing economies such as Nigeria. Through demand curve analysis, the study seeks to understand consumer decision-making patterns and demand elasticity within the Nigerian health insurance market; (2) To explore the impact of market structures, open versus closed economies, on consumer behavior and insurance uptake; (3) To examine how health insurance demand varies across demographic factors including age, gender, and income. This analysis will identify key barriers and motivations affecting insurance demand, providing insights to inform policies aimed at improving coverage accessibility and uptake in the evolving healthcare landscape of developing economies such as Nigeria.

## 2 Study 1

The purpose of Study 1 was to investigate the likelihood of purchasing health insurance in Nigeria, considering price points. To this end, participants were randomly assigned into 2 experimental groups, Open Economy and Closed Economy. The economy is “closed” when health insurance is only available from one source within the decision-making context while “open” economies allow supplementary access to health insurance.

### 2.1 Participants

The study sample consisted of 106 individuals who were recruited from both social media and convenient samples. Participants were included in the study if they were Nigerian citizens and older than 18 years.

### 2.2 Procedure

The Institutional Review Board (IRB) at a State University in the Midwest approved all study procedures. Participants chose to voluntarily engage in this study by clicking on a link which directed them to a web survey hosted by Qualtrics. The first page of the survey presented an informed consent form, and participants continued if they agreed to participate in the study.

#### 2.2.1 Measures

##### Health Insurance Purchase Task (HIPT)

This is a simulated purchase task adapted from Hursh et al. (2020) and was used to assess the likelihood of purchasing health insurance at different price points in Nigerian Naira (₦). At the time of data collection, the exchange rate was approximately ₦1,500 per US Dollar.

Participants were randomly assigned to one of two experimental conditions that differed in their instructions regarding access to health insurance alternatives. All participants received the following base instructions:

> *In the questions that follow we would like you to pretend to purchase 1 month of health insurance coverage with no copay. [CONDITION-SPECIFIC TEXT]. Also, assume that the health insurance coverage you are about to purchase is for your consumption only. In other words, you can’t sell it or give it to anyone else. Everything you buy is, therefore, for your own personal consumption. Please answer the following questions honestly and thoughtfully*.

For participants in the Open Economy condition, the condition-specific text was: *“Imagine that your access to health insurance is not limited to this study and you still have access to your current health insurance plan if you are currently enrolled*.*”* For participants in the Closed Economy condition, the condition-specific text was: *“Assume that the only way you can access health insurance is by purchasing it here. Imagine that any current access you have to health insurance is no longer available and that you are not able to access health insurance outside of this study*.*”*

Participants were then asked a series of dichotomous (Yes/No) questions to indicate their willingness to purchase one month of health insurance with no co-pay at various price points ranging from ₦5.00 to ₦560000.00. The specific price points were: ₦5.00, ₦25.00, ₦65.00, ₦125.00, ₦250.00, ₦500.00, ₦1500.00, ₦3000.00, ₦5500.00, ₦17500.00, ₦35000.00, ₦70000.00, ₦140000.00, ₦280000.00, and ₦560000.00. All 15 price points were presented on a single page, allowing participants to see the full range of prices and adjust their responses accordingly.

##### Demographic Characteristics

Participants reported their gender identity, selecting from the options: *‘Man’, ‘Woman’, ‘Non-binary’*, or *‘I prefer not to answer’*. Age was reported in years. Political views were rated on a visual analog scale (VAS) from 0 (*“Very conservative/not liberal at all”*) to 100 (*“Very liberal/not conservative at all”*). Religiosity or spirituality was similarly rated on a VAS from 0 (*“Not religious/spiritual at all”*) to 100 (*“Extremely religious/spiritual”*). Total annual household income was categorized using four ordinal levels: *‘We have a hard time buying the things we need’, ‘We have just enough money’, ‘We have no problem buying things and sometimes we buy special things’, ‘We have enough money to buy pretty much anything we want’*.

##### Behavioral Measures

This included self-reported use of nicotine, alcohol, marijuana, and gambling behavior. For each behavior, participants indicated *‘Yes’, ‘No’*, or *‘I prefer not to answer’* in response to the following queries: *“Have you ever consumed nicotine products?”, “Have you ever consumed alcohol?”, “Have you ever consumed marijuana?”*, and *“Have you ever gambled?”*

##### Current Health Insurance Status

They reported their health insurance status, choosing from options including *‘Yes, I have partial coverage’, ‘Yes, I have full coverage’, ‘No, I have no health insurance’, ‘No, I am covered through some form of universal healthcare’*, and *‘I prefer not to answer’*.

### 2.3 Statistical Analyses

Exponential demand curves were fit to aggregate data using the model developed by Koffarnus et al. (2015):

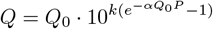

where *Q* represents the percentage of participants purchasing health insurance at a given price point, *Q*_0_ denotes the percentage of participants demanding health insurance when provided at no cost (demand intensity), *α* quantifies the rate of demand decay as price increases (price sensitivity), *P* represents the price level, and *k* serves as a scaling constant reflecting the range of consumption data in log units.

To identify the price point at which demand transitions from inelastic to elastic (*P*_max_), we calculated the price where the first-order derivative of the demand function equals −1, using the Lambert *W* function:

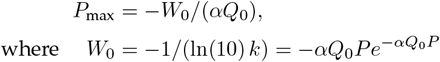

Separate exponential demand models were fitted for open and closed economy conditions and also stratified by demographic and behavioral characteristics to assess parameter stability across subgroups. Model goodness-of-fit was evaluated using *R*^2^ coefficients, with higher values indicating superior model performance. Sensitivity analyses examined the influence of economy type and other participant characteristics on core demand parameters (*Q*_0_ and *α*). Group differences were assessed by comparing model-derived parameter estimates and associated *F*-statistics and *p*-values from the model output. Analyses examined differences across economy type (open vs. closed), age group (above vs. below median), income level (high vs. low), gender (male vs. female), current insurance status (insured vs. uninsured), religiosity, political ideology, and risk behaviors (gambling, cannabis use, nicotine use).

Statistical significance was established at *p <* 0.05 for all analyses. All statistical analyses were conducted using R Statistical Software version 4.4.0 (R Core Team, 2024).

### 2.4 Results

Descriptive statistics for the sample (*N* = 106) are presented in Table 1. Participants had a mean age of *M* = 29.48 years (*SD* = 4.80). Twenty-nine participants (27.36%) identified as male, while 22 participants (20.75%) identified as female. Participants reported moderate to high levels of religiosity (*M* = 77.7, *SD* = 23.00) and moderate levels of conservatism (*M* = 57.60, *SD* = 23.86). Regarding household income, approximately one-quarter of participants (28.21%) reported having “just enough money,” while 14 participants (17.95%) indicated they “have a hard time buying the things we need.” A smaller proportion (11.54%) reported having “no problem buying things and sometimes we buy special things,” and very few participants (1.28%) indicated they “have enough money to buy pretty much anything we want.” Incomplete or missing entries were identified in each category as “Did not respond” (28.3% of the dataset).

**Table 1.** Characteristics of Survey Participants.

| **Variable** | **Mean (SD); n (%)** |
| --- | --- |
| Age | 29.48 (4.80) |
| *Gender* |  |
| Male | 29 (27.36%) |
| Female | 22 (20.75%) |
| Did not respond | 55 (51.89%) |
| *Religiosity* | 77.7 (23.00) |
| Conservatism | 57.60 (23.86) |
| *Household Income* |  |
| We have just enough money | 22 (28.21%) |
| We have a hard time buying the things we need | 14 (17.95%) |
| We have no problem buying things and sometimes we buy special things | 9 (11.54%) |
| We have enough money to buy pretty much anything we want | 1 (1.28%) |
| Did not respond | 32 (54.72%) |
| *Economy Type Assigned To* |  |
| Open | 37 (47.44%) |
| Closed | 41 (52.56%) |
| *Alcohol Consumption* |  |
| No | 33 (31.13%) |
| Yes | 12 (11.32%) |
| I prefer not to answer | 3 (2.83%) |
| Did not respond | 58 (54.72%) |
| *Cannabis Consumption* |  |
| No | 44 (41.51%) |
| Yes | 3 (2.83%) |
| I prefer not to answer | 0 (0%) |
| Did not respond | 59 (55.66%) |
| *Gambling* |  |
| No | 38 (35.85%) |
| Yes | 9 (8.49%) |
| I prefer not to answer | 2 (1.89%) |
| Did not respond | 57 (53.77%) |
| *Nicotine Consumption* |  |
| No | 44 (41.51%) |
| Yes | 4 (3.77%) |
| I prefer not to answer | 0 (0%) |
| Did not respond | 58 (54.72%) |
| *Health Insurance Status* |  |
| Yes, I have full coverage | 3 (2.83%) |
| Yes, I have partial coverage | 8 (7.55%) |
| No, I am covered through some form of universal healthcare | 4 (3.77%) |
| No, I have no health insurance | 31 (29.25%) |
| Did not respond | 60 (56.60%) |

The exponential demand model mostly demonstrated strong predictive validity across various demographic and behavioral factors, with *R*^2^ values ranging from 0.36 to 0.97, as shown in Table 2. The model fits were particularly robust for several subgroups, including participants with low income (*R*^2^ = 0.96), those above median age (*R*^2^ = 0.97), and those without current insurance coverage (*R*^2^ = 0.95). Figure 1 illustrates the aggregate demand curve for health insurance showing how demand varied across different price points.

**Table 2.** Summary Demand Indices Estimated from Aggregate Responses from Nigerian Participants.

| | $Q_0$ | | $\alpha$ | | $P_{\max}$ (₦) | $R^2$ |
| --- | --- | --- | --- | --- | --- | --- |
|  | Estimate | SE | Estimate | SE |  |  |
| *Economy* |  |  |  |  |  |  |
| Open | 76.28 | 3.76 | $5.74 \times 10^{-8}$ | $1.43 \times 10^{-8}$ | 19643.99 | 0.90 |
| Closed | 76.27 | 5.25 | $5.76 \times 10^{-8}$ | $2.01 \times 10^{-8}$ | 19572.98 | 0.74 |
| *Income* |  |  |  |  |  |  |
| High | 74.29 | 3.89 | $3.61 \times 10^{-8}$ | $9.54 \times 10^{-9}$ | 32037.31 | 0.84 |
| Low | 91.37 | 3.39 | $1.65 \times 10^{-7}$ | $2.75 \times 10^{-8}$ | 5710.85 | 0.96 |
| *Age* |  |  |  |  |  |  |
| ≤ Median | 86.31 | 2.76 | $1.45 \times 10^{-7}$ | $2.17 \times 10^{-8}$ | 74092.40 | 0.68 |
| $>$ Median | 72.49 | 4.57 | $1.60 \times 10^{-8}$ | $5.22 \times 10^{-9}$ | 6859.30 | 0.97 |
| *Gender* |  |  |  |  |  |  |
| Male | 75.10 | 3.48 | $2.61 \times 10^{-8}$ | $6.12 \times 10^{-9}$ | 44003.85 | 0.86 |
| Female | 85.44 | 4.38 | $1.64 \times 10^{-7}$ | $3.84 \times 10^{-8}$ | 6141.33 | 0.91 |
| *Religiosity* |  |  |  |  |  |  |
| $>$ Median | 81.92 | 4.06 | $4.34 \times 10^{-8}$ | $1.16 \times 10^{-8}$ | 22646.12 | 0.88 |
| ≤ Median | 72.42 | 4.34 | $6.34 \times 10^{-8}$ | $1.93 \times 10^{-8}$ | 18691.82 | 0.83 |
| *Political Ideology* |  |  |  |  |  |  |
| Conservatism $>$ Median | 75.89 | 4.12 | $4.76 \times 10^{-8}$ | $1.31 \times 10^{-8}$ | 23800.26 | 0.86 |
| Conservatism ≤ Median | 74.21 | 4.81 | $2.99 \times 10^{-8}$ | $9.78 \times 10^{-9}$ | 38812.76 | 0.75 |
| *Currently Insured* |  |  |  |  |  |  |
| Yes | 63.70 | 5.77 | $4.28 \times 10^{-8}$ | $1.95 \times 10^{-8}$ | 31540.85 | 0.53 |
| No | 84.02 | 2.94 | $5.36 \times 10^{-8}$ | $9.48 \times 10^{-9}$ | 19094.11 | 0.95 |
| *Alcohol Use* |  |  |  |  |  |  |
| Yes | 81.43 | 3.50 | $3.73 \times 10^{-8}$ | $8.10 \times 10^{-9}$ | 28323.06 | 0.93 |
| No | 76.12 | 3.37 | $6.38 \times 10^{-8}$ | $1.43 \times 10^{-8}$ | 17717.58 | 0.91 |
| *Cannabis Use* |  |  |  |  |  |  |
| Yes | NC | NC | NC | NC | NC | NC |
| No | 79.02 | 4.45 | $8.52 \times 10^{-8}$ | $2.42 \times 10^{-8}$ | 12775.57 | 0.86 |
| *Gambling* |  |  |  |  |  |  |
| Yes | 69.33 | 5.88 | $3.75 \times 10^{-9}$ | $1.84 \times 10^{-9}$ | 330834.20 | 0.36 |
| No | 77.75 | 3.26 | $7.52 \times 10^{-8}$ | $1.59 \times 10^{-8}$ | 14713.79 | 0.93 |
| *Nicotine Use* |  |  |  |  |  |  |
| Yes | NC | NC | NC | NC | NC | NC |
| No | 76.30 | 4.00 | $5.87 \times 10^{-8}$ | $1.56 \times 10^{-8}$ | 19210.84 | 0.87 |

*Note.* $Q_0$ = demand intensity; $\alpha$ = price sensitivity parameter; $P_{\max}$ = price point estimating where demand transitions from inelastic to elastic; $R^2$ = model goodness-of-fit; $SE$ = standard error. Median splits: age = 29 years, religiosity = 81, conservatism = 60. NC = non-convergence with exponential demand model due to insufficient data. All monetary values are in Nigerian Naira (₦). Higher $\alpha$ values indicate greater price sensitivity (steeper demand decline with price increases).

**Figure 1.**
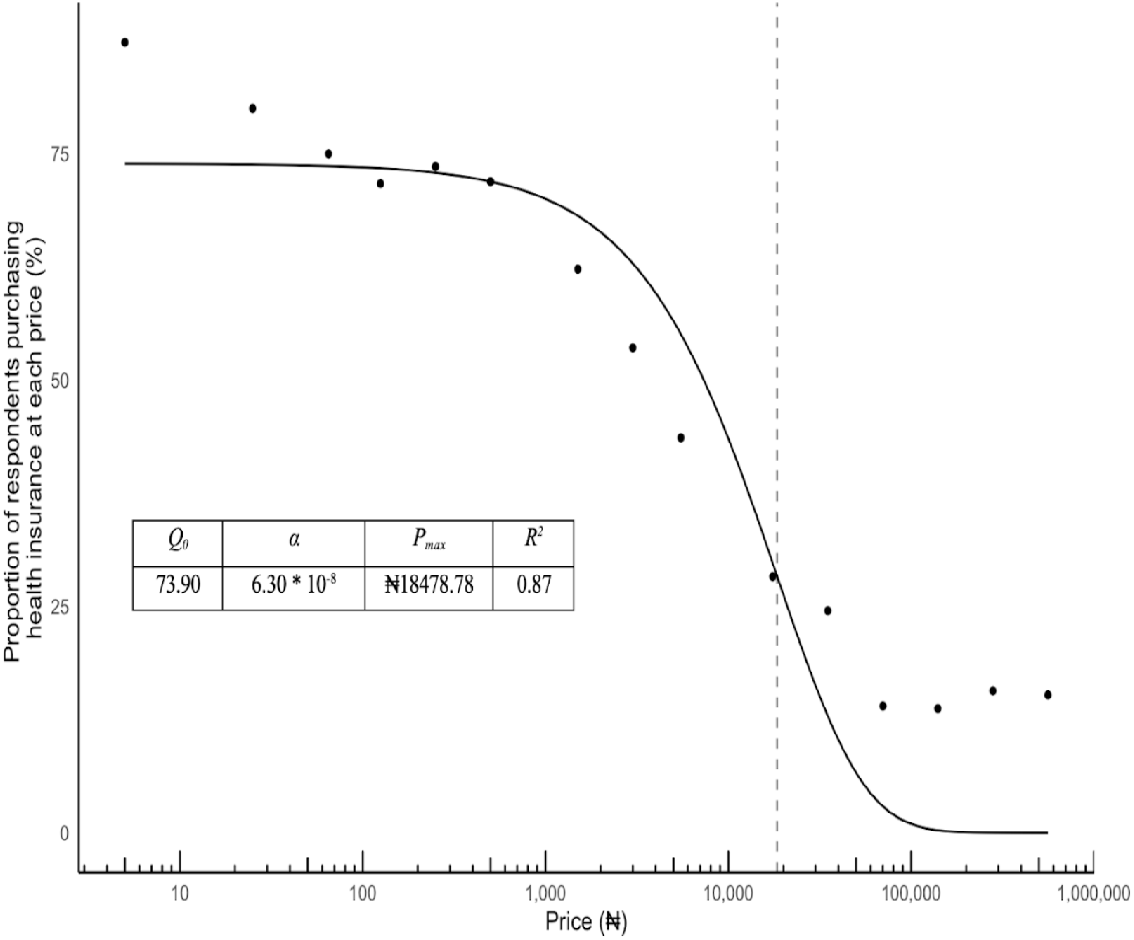
Aggregate Demand for Health Insurance among All Nigerian Respondents. *Note*. Data points represent observed proportions of affirmative responses at each price level. The curve demonstrates typical exponential demand decay, with high initial demand declining as price increases. *Note*. All prices shown in Nigerian Naira (₦). Model fit: *R*^2^ = 0.87, indicating good predictive validity.

**Figure 2.**
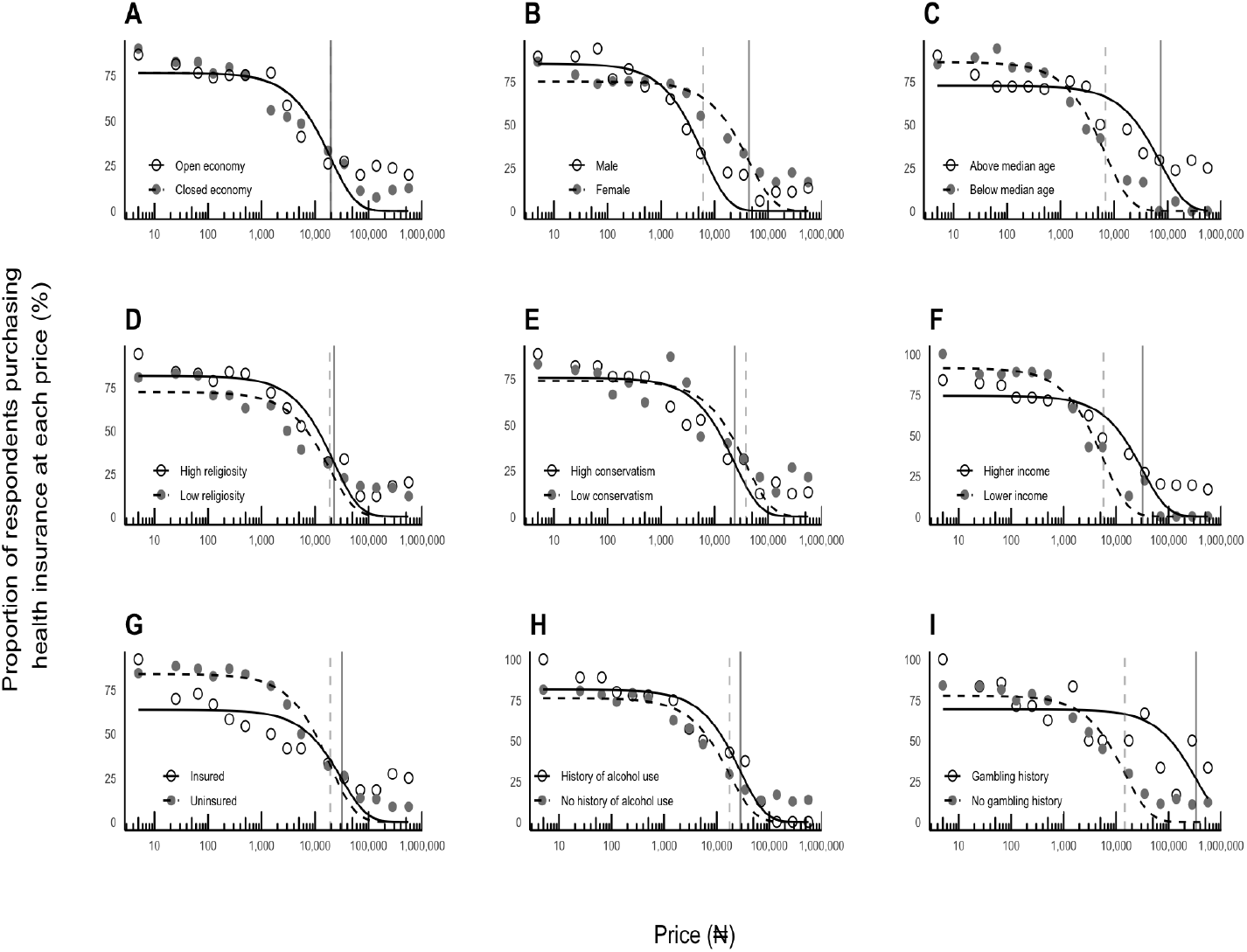
Aggregate Demand for Health Insurance among Subgroups of Respondents. *Note*. Each panel shows the proportion of respondents willing to purchase health insurance at different price points, with fitted exponential demand curves. Panel A: Economy type (open vs. closed economy); Panel B: Gender (male vs. female); Panel C: Age group (above vs. below median age); Panel D: Religiosity level (high vs. low religiosity); Panel E: Political orientation (high vs. low conservatism); Panel F: Income level (higher vs. lower income); Panel G: Insurance status (currently insured vs. uninsured); Panel H: Alcohol use history (history vs. no history of alcohol use); Panel I: Gambling history (gambling history vs. no gambling history). Solid lines and open circles represent the first group in each comparison, while dashed lines and closed circles represent the second group. Vertical lines indicate the price point estimating where demand transitions from inelastic to elastic (*P*_max_) where calculable.

The analysis revealed no significant differences between open and closed economies in terms of either demand intensity (*Q*_0_) (*F* (1, 26) = 0.00, *p* = 0.995) or price sensitivity (*α*) (*F* (1, 26) = 0.00, *p* = 0.999). As detailed in Table 2, both economy types exhibited similar demand patterns, with nearly identical *Q*_0_ values (Open: 76.28, *SE* = 3.76; Closed: 76.27, *SE* = 5.25) and *α* values (Open: 5.74 *×* 10^−8^, *SE* = 1.43 *×* 10^−8^; Closed: 5.76 *×* 10^−8^, *SE* = 2.01 *×* 10^−8^).

The sensitivity analyses presented in Table 3 summarize the effects of several significant demographic factors on demand parameters. Age emerged as a significant factor, with differences observed in both demand intensity (*F* (1, 26) = 5.11, *p* = 0.03) and price sensitivity (*F* (1, 26) = 19.73, *p <* 0.001). Younger participants (below median age) demonstrated higher demand intensity (*Q*_0_ = 86.31, *SE* = 2.76) compared to older participants (*Q*_0_ = 72.49, *SE* = 4.57). Income level similarly influenced both demand intensity (*F* (1, 26) = 9.52, *p* = 0.01) and price sensitivity (*F* (1, 26) = 13.71, *p* = 0.001), with low-income participants showing higher demand intensity (*Q*_0_ = 91.37, *SE* = 3.39) than their high-income counterparts (*Q*_0_ = 74.29, *SE* = 3.89).

**Table 3.** Sensitivity Analyses Examining Patterns of Demand for Health Insurance by Economy Type, Demographics, Behavioral Factors, and Personal Beliefs.

| **Group** | **Parameter** | **F** | **p** |
| --- | --- | --- | --- |
| Economy (Open/Closed) | $Q_0$ | 0.00 | .995 |
| | $\alpha$ | 0.00 | .999 |
| Gender (Male/Female) | $Q_0$ | 3.06 | .09 |
| | $\alpha$ | 15.19 | .001* |
| Age (Above/Below Median) | $Q_0$ | 5.11 | .03* |
| | $\alpha$ | 19.73 | < .001* |
| Income (Above/Below Median) | $Q_0$ | 9.52 | .01* |
| | $\alpha$ | 13.71 | .001* |
| Religiosity (Above/Below Median) | $Q_0$ | 2.19 | .15 |
| | $\alpha$ | 0.33 | .57 |
| Political Ideology (Above/Below Median) | $Q_0$ | 0.06 | .80 |
| | $\alpha$ | 0.70 | .41 |
| Health Insurance (Yes/No) | $Q_0$ | 8.20 | .01* |
| | $\alpha$ | 0.12 | .74 |
| Alcohol Use (Yes/No) | $Q_0$ | 1.11 | .30 |
| | $\alpha$ | 1.94 | .18 |
| Cannabis Use (Yes/No) | $Q_0$ | NA | NA |
| | $\alpha$ | NA | NA |
| Gambling (Yes/No) | $Q_0$ | NA | NA |
| | $\alpha$ | 14.34 | .001* |
| Nicotine Use (Yes/No) | $Q_0$ | NA | NA |
| | $\alpha$ | NA | NA |

*Note.* $F$ -statistics and $p$ -values derived from exponential demand model comparisons between subgroups. $Q_0$ = demand intensity; $\alpha$ = price sensitivity parameter. Median splits: age = 29 years, religiosity = 81, conservatism = 60. Significance threshold set at $p < 0.05$ . NA = not applicable due to model non-convergence for one or more subgroups, preventing statistical comparisons. Bold values indicate statistically significant differences between groups.

Gender differences manifested primarily in price sensitivity (*F* (1, 26) = 15.192, *p* = 0.001) rather than demand intensity (*F* (1, 26) = 3.06, *p* = 0.09). Female participants exhibited higher demand intensity (*Q*_0_ = 85.44, *SE* = 4.38) compared to males (*Q*_0_ = 75.10, *SE* = 3.48). Current insurance status significantly affected demand intensity (*F* = 8.20, *p* = 0.01) but not price sensitivity (*F* (1, 26) = 0.12, *p* = 0.74), with uninsured participants showing notably higher initial demand (*Q*_0_ = 84.02, *SE* = 2.94) than those currently insured (*Q*_0_ = 63.70, *SE* = 5.77).

Personal beliefs, including religiosity and political ideology, showed no significant effects on demand parameters. Religiosity demonstrated no significant differences in either demand intensity (*F* (1, 26) = 2.19, *p* = 0.15) or price sensitivity (*F* (1, 26) = 0.33, *p* = 0.57). Similarly, conservative ideology showed no significant effects on demand intensity (*F* (1, 26) = 0.06, *p* = 0.80) or price sensitivity (*F* (1, 26) = 0.70, *p* = 0.41). Among risk behaviors, gambling behavior showed significant differences in price sensitivity (*F* (1, 26) = 14.34, *p* = 0.001), though demand intensity comparisons could not be computed. Analyses for cannabis and nicotine use were limited due to non-convergence of the demand model for these groups.

Individual *Q*_0_ and *α* parameters could not be estimated due to participants providing dichotomous Yes–No responses at the different price points in the hypothetical purchase task. This response format made it challenging to fit the demand function to individual-level data. A nonparametric Area Under the Curve (AUC) approach (Myerson et al., 2001) was therefore used as a proxy demand index for individual participants. Figure 3 provides a graphical representation of the aggregated normalized AUC values segregated by different experimental or demographic factors. That said, further analyses revealed that neither economy type nor any of the personal/behavioral factors significantly predicted variation in AUC values.

**Figure 3.**
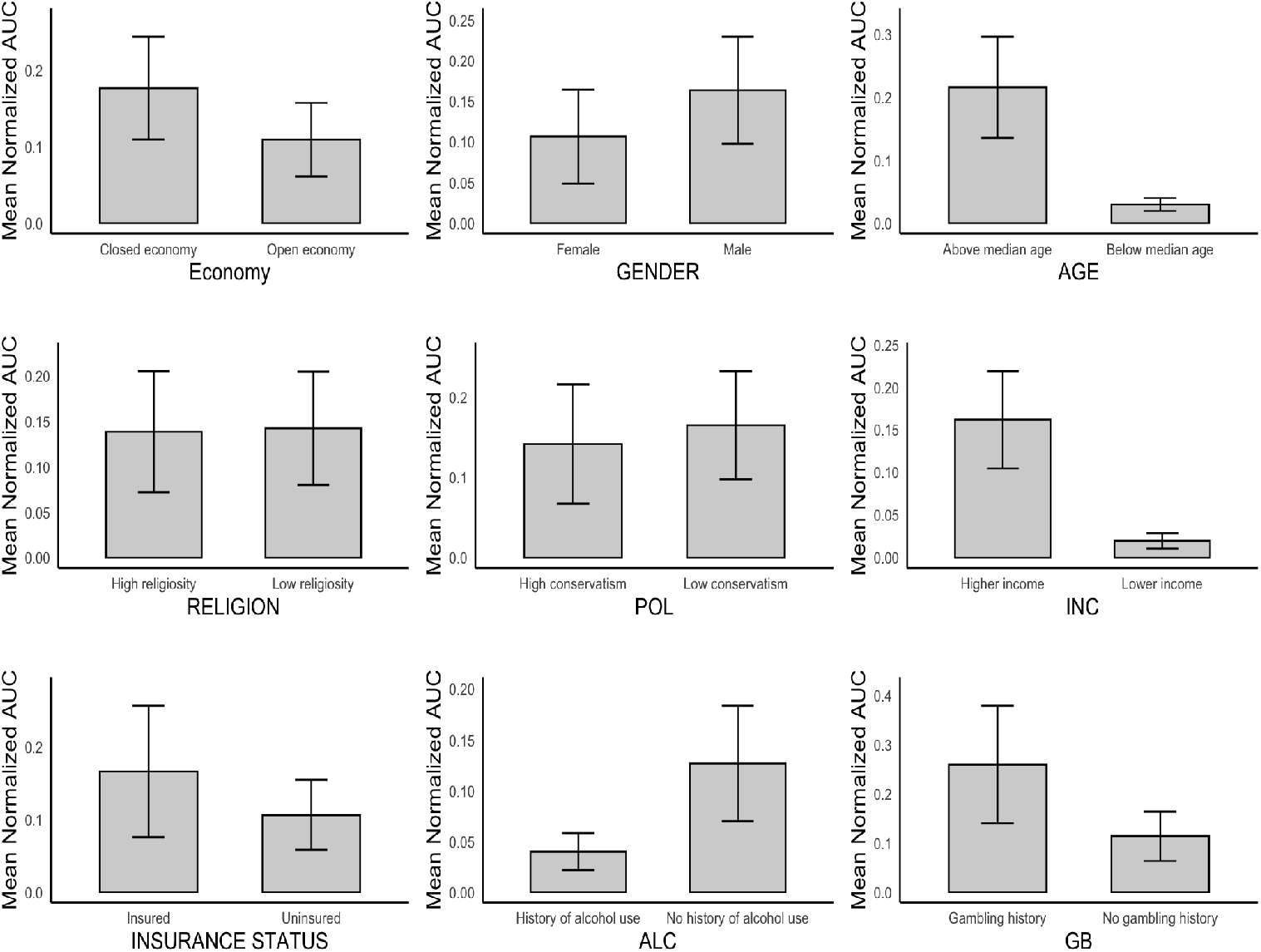
Aggregate Normalized AUCs for Health Insurance among Subgroups of Respondents. *Note*. AUC = Area Under the Curve, a nonparametric measure of overall demand calculated from individual participant responses to the Health Insurance Purchase Task. Values are normalized (0– 1 scale) where higher values indicate greater overall demand for health insurance across all price points. Error bars represent standard error of the mean. No statistically significant differences were observed between any of the compared groups (all *p >* 0.05).

**Figure 4.**
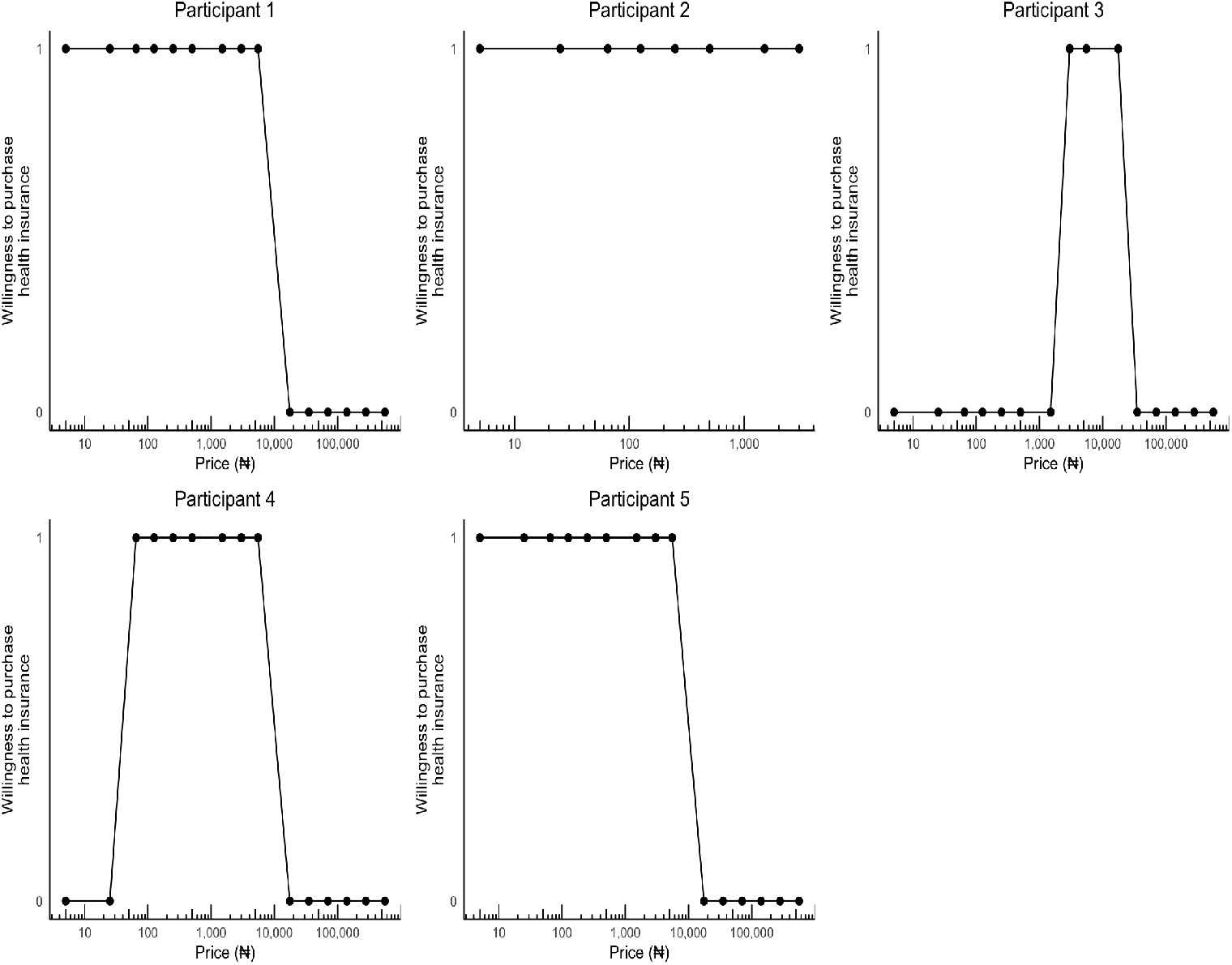
Health Insurance Demand Curves of Focus Group Participants (N = 5)

## 3 Study 2

### 3.1 Methodology

#### 3.1.1 Research Design

Study 2 employed a qualitative research design using focus group discussions (FGDs) to explore in-depth perspectives on health insurance demand in Nigeria. This approach was selected to complement the quantitative findings from Study 1 and to capture nuanced contextual factors influencing health insurance decisions that might not be apparent in the quantitative analysis (Creswell & Poth, 2018).

#### 3.1.2 Participants

Participants were recruited through a non-probability convenience volunteer sampling, whereby they self-selected to participate in the focus group discussions (Etikan et al., 2016; Jager et al., 2017). At the conclusion of Study 1, all participants were invited to indicate their interest in participating in follow-up focus group discussions through a ‘Future Considerations’ section in the survey. This sampling approach acknowledges both the convenience of recruiting participants from an existing sample, and the volunteer element, whereby they self-select, which is appropriate for exploratory qualitative research where depth of insight is prioritized over statistical representativeness (Robinson, 2014). While 12 participants initially expressed interest, 5 individuals (4 females and 1 males) ultimately participated in the focus group discussion. The inclusion criteria remained consistent with Study 1, that is, Nigerian citizenship and aged 18 years or older.

#### 3.1.3 Data Collection

The focus group discussion was conducted approximately two months after the completion of Study 1, in July 2024. Given logistical constraints and geographical dispersion of participants, the FGD was conducted virtually using the Zoom video conferencing platform, which has been established as an effective tool for qualitative data collection (Gray et al., 2020). The session lasted approximately 90 minutes and was facilitated by the lead researcher who is trained in qualitative data collection with experience in behavioral economics research.

The FGD began with participants completing the Health Insurance Purchase Task (HIPT), identical to the one used in Study 1. Only after completing this quantitative measure did participants proceed to the qualitative discussion phase of the focus group. This sequencing was intentional, allowing participants to first engage with concrete pricing decisions before reflecting on their decision-making processes in the subsequent discussion.

Following the HIPT completion, the moderator facilitated a semi-structured discussion based on an interview guide developed from the preliminary findings of Study 1 and the behavioral economic framework underpinning the research. The guide explored several key discussion areas, such as current health insurance status and sources of coverage, perceived value of health insurance for Nigerians, factors influencing health insurance purchase decisions, perceptions of open versus closed health insurance markets, attitudes toward potential government mandates for health insurance, and characteristics of an ideal health insurance package. The focus group was audio-recorded and transcribed verbatim, with participants’ permission.

#### 3.1.4 Data Analyses

The qualitative data for Study 2 were analyzed using thematic analysis (Saunders et al., 2023). Participants’ complete responses were transcribed verbatim and coded accordingly. This systematic process was selected for its rigorous yet flexible framework that accommodates both inductive and deductive analytical approaches (Azungah, 2018). To enhance rigor, both the lead researcher and a junior researcher independently coded the data, then met to compare their coding frameworks, discuss discrepancies, and reach consensus on a unified coding scheme. Next came theme development, where reconciled codes were grouped into potential themes and subthemes. During theme review, identified themes were examined for internal homogeneity and external heterogeneity, ensuring they formed coherent patterns and accurately represented the dataset. The fifth phase involved theme definition and naming, where clear definitions and names were developed for each theme and subtheme. Finally, the report production phase synthesized the analysis into a coherent narrative, with illustrative quotes selected to support key findings. In addition to conducting thematic analysis, the lead researcher derived demand curves to explore each participant’s responses to the HIPT. A table was also created to summarize the participants’ characteristics.

### 3.2 Results

#### 3.2.1 Participant Characteristics

The focus group comprised five participants aged 23-29 years, with four females and one male. Table 4 presents their demographic characteristics.

**Table 4.** Characteristics of Nigerian Focus Group Participants (N = 5)

| Participant | Gender | Subjective Household Income |
| --- | --- | --- |
| 1 | Female | We have a hard time buying the things we need |
| 2 | Female | We have a hard time buying the things we need |
| 3 | Male | We have just enough money |
| 4 | Female | We have just enough money |
| 5 | Female | Not provided |

#### 3.2.2 Thematic Analysis

Six overarching themes emerged from the analysis, each with multiple subthemes. These themes capture the nuances surrounding demand for health insurance in Nigeria.

#### 3.2.3 Sources of health insurance coverage

##### Employer-Provided Coverage

Participants identified employment as a primary avenue for obtaining health insurance. One female participant described her experience: *“I remember when I was working in a bank, so, we had health insurance, and we had a list of hospitals we could visit under this health insurance*.*”* This form of coverage was characterized by restricted provider networks but offered consistent access to care. Participants also noted that university staff, through their employers, had continuous access to healthcare at the school clinics even during breaks. As one participant shared, *“The clinic is not just for students, it’s for staff, so they don’t take breaks like that, they are always working*.*”*

##### Education Institution Coverage

Student status emerged as another pathway to health insurance, with mandatory health fees incorporated into tuition payments. A participant explained: *“We have a university clinic that once you register, you get free access to healthcare* … *There is* [sic] *some health fees we pay. Actually it’s included in our school fees*.*”* This coverage remained active even during academic breaks, providing continuity of care: *“Well, I guess they use* [sic] *to attend to us during the break, because most of the time, they don’t ask for ID card. They just ask for our card numbers when we go there for treatment. So, I guess even if the session is on break* … *they should still attend to us. But I don’t think I’ve ever gone there during the break before, but I feel like if you go there during the break, they will still attend to you, because I’ve never heard people complain that during the break, they were not treated or attended to*.*”*

##### Family-Based Coverage

Participants described accessing insurance through family connections, specifically through the National Health Insurance Scheme (NHIS). This coverage was noted to have age limitations: *“Okay, well, I used to have* [health insurance] *under my parents but when I became eighteen, I had to* … *I’m no longer a dependent on my parents* … *“* Another participant confirmed: *“When I was younger, I was enrolled under NHIS with my dad, and when I grew over eighteen, I have been on my own, so I have to pay out of pocket, ever since*.*”* This finding highlights a crucial point where there is a shift in young adults’ insurance coverage preferences.

#### 3.2.4 Perceived Value of Health Insurance Financial Protection Value

Participants consistently emphasized the financial protection value of health insurance, particularly in the context of Nigeria’s economic challenges. One participant noted: *“Yes, for me, there’s value* … *because it really helps a lot, especially the cost. So, it helps to break down costs, reduce costs and for emergencies too*.*”* Another participant highlighted the impact of inflation on healthcare expenses, making insurance more valuable: *“I think, yes* [there is value]… *And, that’s because* [of] *the inflation of prices of things in the country. Right now, I think I bought malaria drugs the last time for four thousand naira. Yes, it’s that bad now, and this is. a malaria drug you can buy at maybe two thousand* [naira] *in 2022. And it’s 2024 now… and it’s about four thousand* [naira], *three thousand five* [hundred naira], *you can still get some for two thousand six hundred* [naira], *but it’s not strong. If you really want to get the good ones, you will spend between three thousand* [naira] *and four thousand* [naira]. *Do you see that? So, the health insurance, so this is something the health insurance really, really, really, minimize*.*”*

##### Access Facilitation Value

Beyond financial protection, participants valued insurance for facilitating smoother access to healthcare services: *“Apart from those that don’t have health insurance, they have to, maybe wait on queues, you have to go here, to pay this, pay that, so it’s always very stressful actually but when you’re on health insurance, the stress is less, time is, it’s not time-consuming, and the expenses is reduced*.*”* A male participant elaborated on how insurance served as a buffer against inefficiencies in the healthcare system: *“The health insurance… helps you to get the medical test on time. Because with the declining state of health facilities within the country, if you don’t… have that kind of covering, you just spend long hours within the clinic, and everybody, nobody seems to be attending to you. So, I think, the health insurance* … *offers some sort of cushion effect, for the way things are in the country, actually*.*”*

##### Contextual Factors Affecting Perceived Value

The perceived value of insurance was moderated by several contextual factors. Some participants devalued insurance due to perceived immunity to illness: *“What happen* [sic] *is, if I feel a little bit off, well, to God’s praise, I rarely fall sick, but if I do, I just, in those, rare cases, I just go to the pharmacist, and, they prescribe something for me, and I use it, and I’m fine*.*”* Additionally, the type of healthcare facility accessible through insurance influenced its perceived value. Participants pointed out that in government-owned hospitals, having health insurance may not necessarily lead to faster service, which could undermine the overall value of the insurance. One participant explained: *“I think the benefit depends on which hospital you are registered with. If it is a public hospital or a tertiary hospital, the length of hours you have to stay on the queue before a doctor attends to you can be disheartening*.*”*

#### 3.2.5 Price Sensitivity Patterns Income-Based Price Thresholds

Participants demonstrated distinct price sensitivity thresholds that corresponded with their economic circumstances. A student participant explained: *“So, for me, I filled yes, up to five thousand five hundred* [naira]. *Then after five thousand five hundred* [naira], *the next option was seventeen thousand five hundred* [naira]. *So, from there, I filled no, my reason is because, I’m a student, and I have limited sources of income. So, paying seventeen thousand five hundred* [naira] *monthly, for health insurance is, that’s way overboard for me*.*”* An unemployed participant expressed similar constraints: *“I filled yes to five thousand five hundred* [naira] *too, then from, seventeen thousand five hundred* [naira], *I stopped. It was no, because right now I’m currently unemployed. So, just an opinion, and I don’t think I’ll be able to afford that monthly*.*”*

##### Reverse Price Sensitivity

Interestingly, some participants demonstrated what we have termed *“reverse price sensitivity,”* where they rejected extremely low-priced insurance options due to quality concerns. One participant explained: *“I actually did not want to pick those ones* [in] *smaller currencies, because I felt* … *the quality of health care that I would be getting, with that amount will not be so great* … *I’ll still prefer to pay a reasonable amount of money, even if it’s subsidized, so that I can be sure that I’m not getting Paracetamol or Vitamin C. I am sure that this is quality health care, not just a figurehead kind of insurance*.*”* Another participant shared: *“I chose no, from five naira to one thousand five hundred* [naira], *because I tried to live in the reality of things within the country, basically. Because I tried to be realistic, and I know that’s quite low, for the situation*.*”* This finding suggests a non-linear relationship between price and demand for health insurance, with both extremely low and extremely high prices potentially reducing demand.

##### Factors Influencing Price Sensitivity

Participants identified several factors that would reduce their price sensitivity, making them willing to pay higher premiums. Some identified family coverage as a key factor, with one noting: *“If it’s going to involve my family, my husband, children, yes*.*”* Comprehensive benefits also influenced willingness to pay: *“If it’s going to involve other medical attention, not just the basic ones. So, if it’s maybe like an operation* … *I don’t mind paying above that amount*.*”* Quality assurance was similarly important: *“For me to pay more for health insurance, I have to be assured of quality of health care*.*”* Economic stability was also mentioned: *“If the economy is more stable, then seventeen* [thousand] *five* [hundred naira] *is not something I should see as too big*.*”*

#### 3.2.6 Ideal Insurance Package Components

Participants emphasized several essential components for an ideal insurance package. Basic diagnostics and treatment were considered fundamental, as one participant explained: *“I feel like insurance should be able to cover, let’s say I have an emergency lab test and diagnosis, it should be able to like, at least run tests and be able to diagnose whatever it is that is wrong with the person* … *and some basic drugs, not too expensive*.*”* Specialized care was also identified as important: *“I think it should also be able to cover, some special needs like dental care, or, eye care, or I can say people that use glasses, and they need to change their glasses from time to time, or people that want to, you know, check their teeth, dental checkup, and then overall checkup*.*”* Additionally, maternal and child health coverage was deemed essential: *“For me, yeah, basic tests. Basic diagnosis, malaria, typhoid. So, all those basic ones. Then like, maternity too should be included for pregnant women, then children also, pediatrics, should be included for children*.*”*

#### 3.2.7 Perspectives on Government Mandates Concerns About System Integrity

Participants expressed considerable skepticism about government-mandated health insurance, citing concerns about Nigeria’s political environment: *“It’s difficult to say some things in Nigeria because of* … *the way the political phase is, the way the political leaders take decisions* … *if the government mandates it, it has its pros and cons, and it’s good that people will get access to health care easily and at a cheaper cost but in the long run because of the way the country is, it could just get corrupted and it becomes something that is not worth it, after all*.*”*

##### Concerns About Implementation Inequities

Participants worried that even with a mandate, access issues might persist due to favoritism: *“It’s a good one but we should not rejoice totally yet, because the consistency can be shaking* [sic]…*because I have enough insurance covered by the government, and then I go to an hospital or an health care center, and then, I’m deprived of it. So, politics can come into place where you have big people coming in, and then they leave the masses to queue and wait for a very long time. Just because I want to get* [healthcare]…*sometimes some people spend the whole day* … *Even in our healthcare facilities here, you see people waiting for a very long time to even see doctors. So, some guys will consider, I wish I had money to go to a private hospital, so that I will get answered immediately*.*”*

##### Potential Public Health Benefits

Despite concerns, participants acknowledged potential benefits of a mandate, particularly in reducing risky health behaviors: *“I believe*, [if] *the government introduces or mandates health insurance for everyone*, [it] *is a very good idea, because it will give everyone down to the grassroots an opportunity to* [access healthcare]… *there are a lot of people that up till now, they’ve given up on hospitals because they feel they cannot afford it* … *So, I believe that it will still be a good idea if it is introduced and mandated*.*”*

#### 3.2.8 Effects of Alternatives and Substitutes

##### Reliance on Alternative Healthcare Options

Due to the financial burden of out-of-pocket payments, participants noted the emergence of substitutes for formal health insurance. Participants referred to Nigerians who preferred self-medication with over-the-counter medications or herbal remedies over seeking professional medical care. This shift in behavior was linked to a lack of perceived necessity for health insurance. One participant highlighted this substitution effect: *“There is a way that when you have health insurance, you are not quick to, just, you know, go over the counter, and self-prescribe. You’d rather want to do a test. You’d want to know exactly what’s going on, especially when you know that it’s not first about cost, then you prioritize your health. Because the reason why a lot of people do self-medicate is because of the high cost of, so, I feel health insurance does a lot to ensure quality health care, for people generally*.*”* Another participant emphasized the reliance on medicinal herbs and out-of-pocket payments as alternatives to health insurance: *“There are a lot of people that up till now. They’ve given up on hospitals because they feel they cannot afford it. Even general hospitals, most of them still take herbs. They go to pharmacists and all that which have their own risk, but mandating it and giving everyone access to healthcare through the health insurance is to give everyone, at least everyone already has that choice*.*”* These findings suggest that over-the-counter medications, pharmacist consultations, self-medication, and herbal remedies function as proceeding substitutes for formal health insurance in the Nigerian context, particularly among those who face financial constraints.

##### Perceived Benefits of an Open Economy

Participants favored a more decentralized health insurance system with multiple providers, citing several advantages. They noted that quality improvement through competition would be a significant advantage: *“If we have varieties of health insurance, I believe that if people see like, this party or this can give me something better, I believe that it will work* … *competition is even good for businesses*.*”* Geographical accessibility was also identified as an important benefit: *“Well, for me, it will be very okay, so that people can be able to choose where they want to go* … *for location purposes, because some* [health providers] *might be nearer to their side, where they stay*.*”*

#### 3.2.9 Individual Demand Patterns

Figure 2 presents individual health insurance demand curves for the five focus group participants. These curves demonstrated considerable heterogeneity in price sensitivity patterns. Participant 3 showed a distinct bitonic pattern, with willingness to purchase only at middle price points (₦5,500-₦35,000), rejecting both extremely low and high prices. Participant 5 exhibited consistent willingness to purchase up to ₦17,500, with a sharp decline thereafter. Participant 4 showed a similar pattern to 3, rejecting extremely low prices and only willing to purchase up to ₦17,500. Participant 1 demonstrated willingness to purchase up to ₦5,500, with complete rejection at higher price points. Participant 2 maintained a willingness to purchase only up to ₦3,000, after which they did not respond to any higher price points.

## 4 General Discussion

This mixed-methods study investigated factors influencing health insurance demand in Nigeria using behavioral economic principles and hypothetical purchase tasks. Study 1 employed quantitative analysis of demand patterns across open and closed economy conditions, while Study 2 used a focus group discussion following completion of a purchase task to explore participants’ decision-making processes and reasoning. The exponential demand model (Koffarnus et al., 2015) fit the data well, with a median *R*^2^ = 0.87 (*IQR* = 0.81–0.92), indicating strong predictive validity across demographic subgroups. These findings also support the use of hypothetical purchase tasks as validated tools for understanding consumer demand in developing economies (Reed et al., 2022; Khan, 2020). Aggregate demand curves revealed typical patterns of decreasing consumption as price increased, consistent with established behavioral economic models.

One of the most informative quantitative findings was the identification of *P*_max_, the price at which demand shifts from inelastic to elastic. For example, the low-income subgroup had a *P*_max_ of ₦5,710 ($3.81 USD), closely matching focus group participants’ stated cutoff of ₦5,500 ($3.67 USD). While *P*_max_ varied widely across subgroups (from ₦6,141 [$4.10 USD] for females to over ₦74,000 [$49.33 USD] for younger participants), this alignment in the low-income group suggests that, for certain populations, behavioral economic estimates can correspond closely to self-reported affordability limits.

Qualitative findings also revealed a “reverse price sensitivity,” where some participants rejected extremely low-priced insurance options due to quality concerns. While the aggregate demand curves from Study 1 did not show a clear drop in purchases at very low prices, Study 2’s focus group discussions clarified why some participants rejected inexpensive options. Participants worried that very cheap insurance would provide inadequate coverage or poor-quality care, with one noting a preference to *“pay a reasonable amount of money* … *so that I can be sure that I’m not getting Paracetamol or Vitamin C*.*”* This finding aligns with research revealing how quality perceptions influence decision-making in health markets (Casabianca et al., 2022) and suggests that insurance pricing must balance affordability with quality signaling, as prices that are too low may actually reduce demand by suggesting poor value.

Taken together, these findings point to a potential “pricing corridor” for low-to middle-income participants (roughly ₦3,000 to ₦17,500; approximately $2 to $12 USD at the time of data collection) where premiums may be high enough to signal quality but still within the range of relatively inelastic demand. This corridor should not be interpreted as universal, given the substantial variation in *P*_max_ across demographic groups, but it offers a useful starting point for designing targeted premium structures in Nigeria’s health insurance market.

Contrary to the theoretical expectation that consumers should respond differently when alternatives are available versus when they are not (Gilroy et al., 2018; Hursh, 2014; Reed et al., 2013), no significant differences emerged between open and closed economy conditions in demand intensity or price sensitivity. Focus group discussions revealed why this may have occurred: participants already operate within a complex ecosystem of healthcare alternatives, regardless of the experimental condition they were assigned. Even when instructed to imagine that formal insurance was their only option (closed condition), participants’ real-world knowledge of alternatives, including self-medication, over-the-counter pharmaceuticals, traditional medicine, and out-of-pocket payments, likely influenced their responses. As one participant noted, many Nigerians have *“given up on hospitals because they feel they cannot afford it”* and instead rely on pharmacists, medicinal herbs, and self-treatment.

Skepticism towards Nigeria’s National Health Insurance Scheme (NHIS), which is a system characterized by inefficient service delivery, inadequate infrastructure, and resource mismanagement (Eze et al., 2024), may have further eroded the salience of the experimental manipulation. This interpretation aligns with a previous study showing that only half of respondents expressed willingness to consider alternatives to the NHIS due to dissatisfaction with service delivery (Kofoworola et al., 2020). Taken together, this finding has important implications for how researchers model choice contexts in developing economies and highlights the need to consider broader institutional and cultural factors when applying behavioral economic frameworks (Kunreuther & Pauly, 2015; Madrian, 2014).

A related qualitative finding was that most participants accessed insurance through institutions rather than individual purchase, consistent with Adekunle and Vincent (2024). Employment-based coverage was common among formal sector workers, while students accessed care through mandatory health fees incorporated into tuition, and NHIS family-based coverage expired at age 18. This creates a critical transition point where coverage lapses, especially for young adults, and suggests that individual market approaches may be less effective than employer-sponsored or government-sponsored programs in achieving broad coverage (Onwujekwe et al., 2011).

Both age and income significantly influenced health insurance demand, revealing important patterns for understanding target populations. Younger participants demonstrated higher demand intensity when insurance was free or low-cost, but also showed greater price sensitivity as costs increased. The results also support findings by Boonen et al. (2016), who noted that younger adults are more sensitive to the price of health insurance, whereas older adults are more sensitive to quality considerations. Recent evidence from Nigeria also shows that young adults will take up coverage when it is available and accessible (Effiong et al., 2025). This, however, differs from literature suggesting that young adults are less likely to purchase insurance due to perceived invulnerability (Cantiello et al., 2015; Nachiappan et al., 2022).

Low-income participants similarly showed higher demand intensity at zero cost compared to their higher-income counterparts, which aligns with findings by Topan et al. (2024), who demonstrated that income level significantly influences willingness to pay for health insurance. This pattern becomes clearer when considering that low-income individuals face the greatest financial vulnerability to health shocks and have the most to gain from financial protection (Erlangga et al., 2019). These individuals often lack robust social support networks and cannot rely on family resources during health crises, making formal insurance coverage particularly valuable as a safety net (Böhnke & Link, 2017; Lubbers et al., 2020). Additionally, low-income participants may be more familiar with the financial burden of out-of-pocket healthcare payments, having experienced these costs directly, leading them to opt for insurance enrollment as a precautionary measure (Anaba et al., 2022). The high demand among younger and lower-income populations suggests that subsidized programs targeting these groups could achieve high uptake rates. This aligns with international evidence on the effectiveness of targeted subsidies in achieving universal health coverage (Schoen et al., 2010; Hussain et al., 2024).

Female participants exhibited higher demand intensity and greater price sensitivity compared to males. This finding aligns with previous research showing that women are more likely to enroll in health insurance (Agyemang-Duah, 2024; Antabe et al., 2025) and may reflect several factors. Women typically have higher healthcare utilization rates throughout their lives (Kaiser Family Foundation, 2021), particularly related to reproductive health, making insurance coverage more immediately valuable. Additionally, women often serve as healthcare decision-makers for their families (Matoff-Stepp et al., 2014), potentially increasing their awareness of healthcare costs and the value of insurance protection.

Participants’ risk-taking behaviors revealed complex relationships with health insurance demand. Those who gambled showed lower price sensitivity than non-gamblers, suggesting that risk-tolerant individuals may be more willing to pay higher premiums. This finding has two possible explanations. First, gambling behavior is associated with various health problems and declining health outcomes (Butler et al., 2020; Jazaeri & Habil, 2012; Latvala et al., 2019), potentially making gamblers more aware of their need for healthcare access. The behavioral tendency toward risk-taking, combined with declining health outcomes among frequent gamblers, may increase their reliance on medical care and consequently their willingness to secure access to treatment, even if it involves higher costs (Dąbrowska et al., 2017). Second, individuals comfortable with financial risk-taking in gambling contexts may view health insurance premiums as an acceptable wager against future health problems, or alternatively, may view health insurance as a way to secure health protection while continuing to engage in risky behaviors (Butler et al., 2020).

Similarly, participants with alcohol consumption history showed higher demand intensity compared to non-users. This pattern likely reflects their increased vulnerability to alcohol-related health conditions and higher expected healthcare utilization, as people with alcohol-related diagnoses incur significantly higher healthcare costs due to conditions such as heart disease, stroke, liver disease, and certain cancers (Ozluk, 2022). The higher demand for health insurance may also be explained by the concept of moral hazard, as discussed by Azagba et al. (2021), who found a significant relationship between health insurance coverage and heavy drinking rates.

Interestingly, religious and political beliefs showed no statistically significant effects on health insurance demand. The lack of religious influence was particularly surprising, given previous research suggesting that faith-based beliefs about divine providence might reduce demand for insurance (Adewole et al., 2015). However, quantitative analyses in the present study revealed that more religious participants actually had higher demand intensity, suggesting that religious individuals may view health insurance as not conflicting with faith-based beliefs. This finding also differs from Özden et al. (2024), whose study found that individuals without specific religious affiliations were more likely to purchase insurance.

Focus group participants valued health insurance primarily for financial protection and improved access to care, particularly in the context of Nigeria’s high inflation and rising healthcare costs, consistent with findings by Erlangga et al. (2019) who emphasized the financial protection offered by public health insurance. As one participant noted, malaria treatment costs had doubled between 2022 and 2024, making insurance coverage increasingly valuable for managing routine healthcare expenses. This finding also aligns with Adams (2024), who highlighted how economic stagnation and social inequalities in Nigeria collectively constrain access to essential healthcare services. That said, value perceptions were tempered by concerns with inefficiencies in the healthcare system. Focus group participants noted that having insurance did not guarantee faster service at government hospitals, a concern that reflects broader challenges across the six World Health Organization health system pillars (Oleribe et al., 2019). These structural issues including delayed care, workforce constraints, medicine shortages, financing gaps, and weak governance, undermine insurance’s perceived benefits, particularly in rural areas (Abah, 2023).

Views on government-mandated health insurance were mixed. While some acknowledged its potential to improve coverage and reduce reliance on unsafe alternatives such as unregulated medications and traditional remedies, others feared that corruption, political interference, and implementation inequities would limit its effectiveness. These concerns echo Bashar et al. (2025), who identified governance, market efficiency, and organizational health as priorities for expanding participation in Nigerian health insurance schemes. The findings suggest that successful insurance expansion requires concurrent improvements in government transparency and accountability to build public trust.

This research makes a significant contribution by demonstrating the successful application of hypothetical purchase tasks (HPTs) to health insurance demand in a developing economy context, extending the validated use of these methods beyond their traditional applications (e.g., Murphy & MacKillop, 2006). The strong model fits in a Nigerian sample support the cross-cultural validity of behavioral economic demand models and demonstrates the value of mixed-methods approaches for understanding health insurance demand. Study 2’s sequential design, where participants completed the hypothetical purchase task before engaging in focus group discussions, allowed for rich reflection on actual choice processes and provided context for interpreting the quantitative patterns observed in Study 1. This approach addresses calls for integrating behavioral economics with qualitative research to better understand decision-making processes (Beatty et al., 2023; Matjasko et al., 2016; Tewogbola et al., 2025).

Several areas warrant further investigation. First, future research should explore how alternative healthcare options (traditional medicine, self-medication, informal networks) function as substitutes for formal insurance in different economic contexts. Second, longitudinal studies could examine how insurance demand changes as individuals transition between institutional coverage types (family to individual, student to employee). Finally, experimental research could test whether the reverse price sensitivity finding replicates in other contexts and commodities, as this has important implications for behavioral economic theory.

The current research has limitations to consider when interpreting its findings. The quantitative component (Study 1) faced challenges related to sample size and data quality. After excluding participants with incomplete or missing responses, 28.3% of the original dataset was removed, reducing the final sample to 76 participants. This high exclusion rate may have introduced selection bias, as participants who completed all survey components may differ systematically from those who provided incomplete responses. Additionally, the use of crowdsourcing for participant recruitment, while practical for accessing Nigerian participants, represents a non-probability sampling method that may not accurately represent the broader Nigerian population, particularly those without reliable internet access or familiarity with online surveys (Cornesse et al., 2020). The binary response format of the hypothetical purchase task, while appropriate for aggregate analysis, prevented the estimation of individual-level demand parameters (*Q*_0_ and *α*), limiting our ability to examine individual differences in demand characteristics. Furthermore, several subgroup analyses were compromised by non-convergence issues, particularly for cannabis and nicotine users, due to insufficient sample sizes in these categories. This prevented comprehensive analysis of substance use effects on health insurance demand.

The qualitative component (Study 2) was limited by its small sample size of five participants, which restricts the depth and generalizability of insights obtained from the focus group discussion (Vasileiou et al., 2018). The use of virtual focus groups via Zoom, while necessary due to geographical constraints, introduced technical challenges including network connectivity issues that occasionally prevented full participant engagement simultaneously (Nehls et al., 2015). Additionally, the participants were predominantly from similar geographic and socioeconomic backgrounds, which may have limited the diversity of perspectives captured and potentially overlooked important regional or cultural variations in health insurance attitudes within Nigeria (Gross et al., 2022). Finally, the study was conducted during a specific economic period in Nigeria, and the findings may not generalize to different economic conditions or policy environments. The rapid inflation mentioned by participants, for example, may have influenced price sensitivity in ways that might not persist under more stable economic conditions.

## Conclusion

This study provides one of the first applications of behavioral economic demand modeling to health insurance in a developing economy. Using hypothetical purchase tasks, we found that exponential demand models effectively characterized health insurance demand in Nigeria. We also uncovered a novel “reverse price sensitivity” where participants systematically rejected extremely low premiums due to quality concerns. Contrary to expectations, no differences emerged between open and closed economy conditions, reflecting the influence of informal healthcare alternatives and institutional distrust that override experimental manipulations. The identification of a critical pricing corridor of ₦3,000 to ₦17,500 ($2 to $12 USD) that balances affordability with quality signaling, combined with evidence that younger and lower-income populations show higher demand intensity but greater price sensitivity, provides actionable insights for designing policies that address both price and system inefficiencies.

## Data Availability

All data produced in the present study are available upon reasonable request to the authors

## Notes

### Competing Interest Statement

The authors have declared no competing interest.

### Funding Statement

This study did not receive any funding.

### Author Declarations

The Institutional Review Board (IRB) of Southern Illinois University gave ethical approval for this work.

## References

Abah, V. O. (2022). Poor health care access in nigeria: A function of fundamental misconceptions and misconstruction of the health system. In Healthcare access - new threats, new approaches [working title]. IntechOpen. 10.5772/intechopen.108530

Abubakar, I., Dalglish, S. L., Angell, B., Sanuade, O., Abimbola, S., Adamu, A. L., Adetifa, I. M. O., Colbourn, T., Ogunlesi, A. O., Onwujekwe, O., Owoaje, E. T., Okeke, I. N., Adeyemo, A., Aliyu, G., Aliyu, M. H., Aliyu, S. H., Ameh, E. A., Archibong, B., Ezeh, A., & Gadanya, M. A. (2022). The lancet nigeria commission: Investing in health and the future of the nation. The Lancet, 399(10330), 1155–1200.

Adams, E. U. (2024). The importance of project in underdeveloped countries. Journal of African Studies and Development, 16(1), 10–24. 10.5897/jasd2023.0692

Adekunle, W., & Vincent, O. (2024). Analysing the determinants of healthcare insurance uptake in nigeria (AERC Working Paper HCD-CCS-009). The Nigerian Economic Summit Group; African Economic Research Consortium. https://publication.aercafricalibrary.org/server/api/core/bitstreams/89750a45-0f13-468a-b8ca-198390ccd503/content

Adeleye, O. A., & Adebamowo, C. A. (2012). Factors associated with research wrongdoing in nigeria. Journal of Empirical Research on Human Research Ethics, 7(5), 15–24. 10.1525/jer.2012.7.5.15

Adewole, D. A., Adebayo, A. M., Udeh, E. I., Shaahu, V. N., & Dairo, M. D. (2015). Payment for health care and perception of the national health insurance scheme in a rural area in south-west nigeria. The American Journal of Tropical Medicine and Hygiene, 93(3), 648–654. 10.4269/ajtmh.14-0245

Agyemang-Duah, W., Oduro, M. S., Peprah, P., Adei, D., & Nkansah, J. O. (2024). The role of gender in health insurance enrollment among geriatric caregivers: Results from the 2022 informal caregiving, health, and healthcare survey in ghana. BMC Public Health, 24(1), 1566. 10.1186/s12889-024-18930-y

Ajike, S., Ezinne, A., & Folarin, M. (2020). Evaluating end users’ knowledge and perceived benefit of the national health insurance scheme post implementation in a south-west state of nigeria. World Journal of Advanced Research and Reviews, 2020(03), 2581–9615. https://pdfs.semanticscholar.org/fda8/e82348ee497bdba44905a5ab18d679f853c2.pdf

Akinyemi, O. O., Owopetu, O. F., & Agbejule, I. O. (2021). National health insurance scheme: Perception and participation of federal civil servants in ibadan. Annals of Ibadan Postgraduate Medicine, 19(1), 49–55.

Allen, R. G. D. (1938). Mathematical analysis for economists. MacMillan & Co., Limited.

Amu, H., Dickson, K. S., Adde, K. S., Kissah-Korsah, K., Darteh, E. K. M., & Kumi-Kyereme, A. (2022). Prevalence and factors associated with health insurance coverage in urban subsaharan africa: Multilevel analyses of demographic and health survey data. PLOS ONE, 17(3), e0264162. 10.1371/journal.pone.0264162

Anaba, E. A., Tandoh, A., Sesay, F. R., & Fokukora, T. (2022). Factors associated with health insurance enrolment among ghanaian children under the five years: Analysis of secondary data from a national survey. BMC Health Services Research, 22(1). 10.1186/s12913-022-07670-7

Antabe, R., Anfaara, F. W., Sano, Y., & Amoak, D. (2025). Ghana’s national health insurance enrollment: Does the intersection of educational and residential status matter? PLOS ONE, 20(2), e0318202.

Anyadighibe, J. (2022). Factors influencing the patronage of insurance companies. Calabar Journal of Finance and Banking, 3(1). https://www.cjobaf.com/pages/Vol3-No1Page133-150.pdf

Aron-Dine, A., Einav, L. E., & Finkelstein, A. (2013). The rand health insurance experiment, three decades later. The Journal of Economic Perspectives, 27(1), 197–222. 10.1257/jep.27.1.197

Azagba, S., Shan, L., Wolfson, M., Hall, M., & Chaloupka, F. (2021). Problem drinking as intentional risky behavior: Examining the association between state health insurance coverage and excessive alcohol consumption. Preventive Medicine Reports, 24, 101556. 10.1016/j.pmedr.2021.101556

Azungah, T. (2018). Qualitative research: Deductive and inductive approaches to data analysis. Qualitative Research Journal, 18(4), 383–400. 10.1108/QRJ-D-18-00035

Bashar, J. M., Hadiza, S., Ugochi, O. J., Muhammad, L. S., Olufemi, A., Eberechi, U., Agada-Amade, Y., Yusuf, A., Abdullahi, A. H., Musa, H. S., Ibrahim, A. A., Nnennaya, K. U., Anyanti, J., Yusuf, D., Okoineme, K., Adebambo, J., Ikani, S. O., Aizobu, D., Abubakar, M., … Wada, Y. H. (2025). Charting the path to the implementation of universal health coverage policy in nigeria through the lens of delphi methodology. BMC Health Services Research, 25(1), 45. 10.1186/s12913-024-12201-7

Beatty, A., Moffitt, R., & Buttenheim, A. (2023). Development of behavioral economics [National Academies Press (US)].

Berry, M. S., Naudé, G. P., Johnson, P. S., & Johnson, M. W. (2023). The blinded-dose purchase task: Assessing hypothetical demand based on cocaine, methamphetamine, and alcohol administration. Psychopharmacology, 240(4), 921–933. 10.1007/s00213-023-06334-6

Böhnke, P., & Link, S. (2017). Poverty and the dynamics of social networks: An analysis of german panel data. European Sociological Review, 33(4), 615–632. 10.1093/esr/jcx063

Boonen, L. H., Laske-Aldershof, T., & Schut, F. T. (2016). Switching health insurers: The role of price, quality and consumer information search. The European Journal of Health Economics, 17(3), 339–353. 10.1007/s10198-015-0681-1

Brown, J., Washington, W. D., Stein, J. S., & Kaplan, B. A. (2021). The gym membership purchase task: Early evidence towards establishment of a novel hypothetical purchase task. The Psychological Record, 72(3), 371–381. 10.1007/s40732-021-00475-w

Butler, N., Quigg, Z., Bates, R., Sayle, M., & Ewart, H. (2020). Gambling with your health: Associations between gambling problem severity and health risk behaviours, health and wellbeing. Journal of Gambling Studies, 36(2), 527–538. 10.1007/s10899019-09902-8

Cantiello, J., Fottler, M. D., Oetjen, D., & Zhang, N. J. (2015). The impact of demographic and perceptual variables on a young adult’s decision to be covered by private health insurance. BMC Health Services Research, 15, 195. 10.1186/s12913-015-0848-6

Casabianca, M. S., Gallego, J. M., Góngora, P., & Rodríguez-Lesmes, P. (2022). Price elasticity of demand for voluntary health insurance plans in colombia. BMC Health Services Research, 22, 618. 10.1186/s12913-022-07899-2

Chukwu, N., Ebue, M., Christy, O., Arionu, N., Patricia, A., & Prince, A. (2016). Problems of social research in nigeria. Research on Humanities and Social Sciences, 6(12), 52–59.

Creswell, J. W., & Poth, C. N. (2016). Qualitative inquiry and research design: Choosing among five approaches. Sage Publications.

Dąbrowska, K., Moskalewicz, J., & Wieczorek, Ł. (2017). Barriers in access to the treatment for people with gambling disorders. are they different from those experienced by people with alcohol and/or drug dependence? Journal of Gambling Studies, 33(2), 487–503. 10.1007/s10899-016-9655-1

Dey, P., & Bach, P. B. (2019). The 6 functions of health insurance. JAMA, 321(13), 1242. 10.1001/jama.2019.2320

Effiong, F. B., Dine, R. D., Hassan, I. A., Olawuyi, D. A., Isong, I. K., & Adewole, D. A. (2025). Coverage and predictors of enrollment in the state-supported health insurance schemes in nigeria: A quantitative multi-site study. BMC Public Health, 25(1), 2125. 10.1186/s12889-025-23329-4

Erlangga, D., Suhrcke, M., Ali, S., & Bloor, K. (2019). The impact of public health insurance on health care utilisation, financial protection and health status in low- and middle-income countries: A systematic review. PLOS ONE, 14(8). 10.1371/journal.pone.0219731

Etikan, I., Musa, S. A., & Alkassim, R. S. (2016). Comparison of convenience sampling and purposive sampling. American Journal of Theoretical and Applied Statistics, 5(1), 1–4.

Eze, O. I., Iseolorunkanmi, A., & Adeloye, D. (2024). The national health insurance scheme (nhis) in nigeria: Current issues and implementation challenges. Journal of Global Health Economics and Policy, 4, e2024002. 10.52872/001c.120197

Gilroy, S. P., Kaplan, B. A., & Reed, D. D. (2020). Interpretation(s) of elasticity in operant demand. Journal of the Experimental Analysis of Behavior, 114(1), 106–115. 10.1002/jeab.610

Gray, L. M., Wong-Wylie, G., Rempel, G. R., & Cook, K. (2020). Expanding qualitative research interviewing strategies: Zoom video communications. The Qualitative Report, 25(5), 1292– 1301.

Gross, A. S., Harry, A. C., Clifton, C. S., & Della Pasqua, O. (2022). Clinical trial diversity: An opportunity for improved insight into the determinants of variability in drug response. British Journal of Clinical Pharmacology, 88(6), 2700–2717. 10.1111/bcp.15242

Hursh, S. R., Strickland, J. C., Schwartz, L. P., & Reed, D. D. (2020). Quantifying the impact of public perceptions on vaccine acceptance using behavioral economics. Frontiers in Public Health, 8. 10.3389/fpubh.2020.608852

Hussain, A., Umair, M., Khan, S., Alonazi, W. B., Almutairi, S. S., & Malik, A. (2024). Exploring sustainable healthcare: Innovations in health economics, social policy, and management. Heliyon, 10(13), e33186. 10.1016/j.heliyon.2024.e33186

Iqlima, N. (2024). Healthcare expenses in the age of covid-19: Strategies for financial preparedness [Available at https://ssrn.com/abstract=4882494].

Jacobs, E. A., & Bickel, W. K. (1999). Modeling drug consumption in the clinic using simulation procedures: Demand for heroin and cigarettes in opioid-dependent outpatients. Experimental and Clinical Psychopharmacology, 7(4), 412.

Jager, J., Putnick, D. L., & Bornstein, M. H. (2017). Ii. more than just convenient: The scientific merits of homogeneous convenience samples. Monographs of the Society for Research in Child Development, 82(2), 13–30.

Jazaeri, S. A., & Habil, M. H. (2012). Reviewing two types of addiction - pathological gambling and substance use. Indian Journal of Psychological Medicine, 34(1), 5–11. 10.4103/0253-7176.96147

Kaiser Family Foundation. (2021). Women’s health care utilization and costs: Findings from the 2020 kff women’s health survey (tech. rep.). Kaiser Family Foundation. https://www.kff.org/womens-health-policy/issue-brief/womens-health-care-utilization-and-costs-findings-from-the-2020-kff-womens-health-survey/

Khan, H. (2020). Behavioral economic analysis of demand for hypothetical work performance: A partial replication. McNair Research Journal SJSU, 16(1), 7. 10.31979/mrj.2020.1607

Kofoworola, A. A., Ekiye, A., Motunrayo, A. O., Adeoye, A. T., & Adunni, M. R. (2020). National health insurance scheme: An assessment of service quality and clients’ dissatisfaction. Ethiopian Journal of Health Sciences, 30(5), 795–802. 10.4314/ejhs.v30i5.20

Kunreuther, H. C., & Pauly, M. V. (2015). Behavioral economics and insurance: Principles and solutions. In D. Schwarcz & P. Siegelman (Eds.), Research handbook on the economics of insurance law (pp. 15–35). Edward Elgar Publishing. 10.4337/9781782547143.00007

Latvala, T., Lintonen, T., & Konu, A. (2019). Public health effects of gambling debate on a conceptual model. BMC Public Health, 19(1), 1077. 10.1186/s12889-019-7391-z

Lubbers, M. J., Small, M. L., & García, H. V. (2020). Do networks help people to manage poverty? perspectives from the field. The Annals of the American Academy of Political and Social Science, 689(1), 7–25. 10.1177/0002716220923959

Madrian, B. C. (2014). Applying insights from behavioral economics to policy design. Annual Review of Economics, 6(1), 663–688. 10.1146/annurev-economics-080213-041033

Matjasko, J. L., Cawley, J. H., Baker-Goering, M. M., & Yokum, D. V. (2016). Applying behavioral economics to public health policy. American Journal of Preventive Medicine, 50(5), S13–S19. 10.1016/j.amepre.2016.02.007

Matoff-Stepp, S., Applebaum, B., Pooler, J., & Kavanagh, E. (2014). Women as health care decisionmakers: Implications for health care coverage in the united states. Journal of Health Care for the Poor and Underserved, 25(4), 1507–1513.

Murphy, J. G., & MacKillop, J. (2006). Relative reinforcing efficacy of alcohol among college student drinkers. Experimental and Clinical Psychopharmacology, 14(2), 219. 10.1037/1064-1297.14.2.219

Myerson, J., Green, L., & Warusawitharana, M. (2001). Area under the curve as a measure of discounting. Journal of the Experimental Analysis of Behavior, 76(2), 235–243. 10.1901/jeab.2001.76-235

Nachiappan, N., Mackinnon, S., Ndayizeye, J. P., Greenfield, G., & Hargreaves, D. (2022). Barriers to accessing health care among young people in 30 low-middle income countries. Health Science Reports, 5(4). 10.1002/hsr2.733

Nehls, K., Smith, B. D., & Schneider, H. A. (2015). Video-conferencing interviews in qualitative research. In S. Hai-Jew (Ed.), Enhancing qualitative and mixed methods research with technology (pp. 140–157). IGI Global Scientific Publishing. 10.4018/978-1-4666-6493-7.ch006

Nigerian Institute of Social and Economic Research. (2024). Aerc/niser policy dialogue on october 10, 2024 - niser [Published October 16, 2024]. https://niser.gov.ng/v2/aerc-niser-policy-dialogue/

Oleribe, O. E., Momoh, J., Uzochukwu, B. S., Mbofana, F., Adebiyi, A., Barbera, T., Williams, R., & Taylor, S. D. (2019). Identifying key challenges facing healthcare systems in africa and potential solutions. International Journal of General Medicine, 12, 395–403. 10.2147/ijgm.s223882

Onwujekwe, O. E., Uzochukwu, B. S., Ezeoke, O. P., & Uguru, N. P. (2011). Health insurance: Principles, models and the nigerian national health insurance scheme. International Journal of Medicine and Health Development, 16(1). https://www.ajol.info/index.php/jcm/article/view/73361

Özden, A. T., Küçükkocaoğlu, G., & Farnoudkia, H. (2024). The impact of religion on insurance purchasing behavior: A dual perspective with economic factors. International Journal of Religion, 5(12), 99–120. 10.61707/41z6cv58

Ozluk, P., Cobb, R., Sylwestrzak, G., Raina, D., & Bailly, E. (2022). Alcohol-attributable medical costs in commercially insured and medicaid populations. AJPM Focus, 1(2), 100036. 10.1016/j.focus.2022.100036

PricewaterhouseCoopers. (2024). Us healthcare consumer insights and engagement survey (tech. rep.). PwC. https://www.pwc.com/us/en/industries/health-industries/library/healthcare-consumer-insights-survey.html

Reed, D. D., Gelino, B. W., & Strickland, J. C. (2022). Behavioral economic demand: How simulated behavioral tasks can inform health policy. Policy Insights from the Behavioral and Brain Sciences, 9(2), 171–178. 10.1177/23727322221118668

Reed, D. D., Niileksela, C. R., & Kaplan, B. A. (2013). Behavioral economics: A tutorial for behavior analysts in practice. Behavior Analysis in Practice, 6(1), 34–54. 10.1007/bf03391790

Robinson, O. C. (2014). Sampling in interview-based qualitative research: A theoretical and practical guide. Qualitative Research in Psychology, 11(1), 25–41.

Sanil, M., & Eminer, F. (2021). An integrative model of patients’ perceived value of healthcare service quality in north cyprus. Archives of Public Health, 79(1). 10.1186/s13690-021-00738-6

Saunders, C. H., Sierpe, A., von Plessen, C., Kennedy, A. M., Leviton, L. C., Bernstein, S. L., Goldwag, J., King, J. R., Marx, C. M., Pogue, J. A., Saunders, R. K., Van Citters, A., Yen, R. W., Elwyn, G., Leyenaar, J. K., & Laboratory, C. (2023). Practical thematic analysis: A guide for multidisciplinary health services research teams engaging in qualitative analysis. BMJ (Clinical research ed.), 381, e074256. 10.1136/bmj-2022-074256

Schoen, C., Osborn, R., Squires, D., Doty, M. M., Pierson, R., & Applebaum, S. (2010). How health insurance design affects access to care and costs, by income, in eleven countries. Health Affairs, 29(12), 2323–2334. 10.1377/hlthaff.2010.0862

Secades-Villa, R., Pericot-Valverde, I., & Weidberg, S. (2016). Relative reinforcing efficacy of cigarettes as a predictor of smoking abstinence among treatment-seeking smokers. Psychopharmacology, 233, 3103–3112. 10.1007/s00213-016-4350-6

Strickland, J. C., Marks, K. R., & Bolin, B. L. (2020). The condom purchase task: A hypothetical demand method for evaluating sexual health decision-making. Journal of the Experimental Analysis of Behavior, 113(2), 435–448. 10.1002/jeab.585

Strickland, J. C., Reed, D. D., Hursh, S. R., Schwartz, L. P., Foster, R. N., Gelino, B. W., & Johnson, M. W. (2022). Behavioral economic methods to inform infectious disease response: Prevention, testing, and vaccination in the covid-19 pandemic. PloS One, 17(1), e0258828. 10.1371/journal.pone.0258828

Tewogbola, P., Jacobs, E. A., Lee, Y. T., Redner, R. N., McDaniel, J. T., & Asirvatham, J. (2025). Beyond the jab: Modeling hiv vaccine acceptance in sexual and gender minorities with behavioral economic demand. Journal of the Experimental Analysis of Behavior, 124(1), e70038.

Topan, G. J., Thiombiano, N., & Sarambe, I. (2024). Determinants of households’ willingness to pay for health insurance in burkina faso. Health Economics Review, 14(1), 1–12. 10.1186/s13561-024-00576-6

Vasileiou, K., Barnett, J., Thorpe, S., & Young, T. (2018). Characterising and justifying sample size sufficiency in interview-based studies: Systematic analysis of qualitative health research over a 15-year period. BMC Medical Research Methodology, 18(1), 148. 10.1186/s12874-018-0594-7

